# Ketone body 3-hydroxybutyrate: a biomarker of aggression?

**DOI:** 10.1101/2020.05.13.20100347

**Authors:** AM Whipp, E Vuoksimaa, T Korhonen, R Pool, RSL Ligthart, FA Hagenbeek, M Bartels, LH Bogl, L Pulkkinen, RJ Rose, DI Boomsma, J Kaprio

## Abstract

Human aggression is a complex behavior, the biological underpinnings of which remain poorly known. To gain insights into aggression biology, we studied relationships with aggression of 11 low-molecular-weight metabolites (amino acids, ketone bodies), processed using ^1^H nuclear magnetic resonance spectroscopy. We used a discovery sample of young adults and an independent adult replication sample. We studied 725 young adults from a population-based Finnish twin cohort born 1983-87, with aggression levels rated in adolescence (ages 12, 14, 17) by multiple raters and blood plasma samples at age 22. Linear regression models specified metabolites as the dependent and aggression ratings as independent variables, and included several fixed effects. All metabolites showed low correlations with aggression, with only one –– 3- hydroxybutyrate, a ketone body produced during fasting –– showing significant (negative) associations with aggression. Effect sizes for different raters were generally similar in magnitude, while teacher-rated (age 12) and self-rated aggression (age 14) were both significant predictors of 3-hydroxybutyrate in multi-rater models. In an independent replication sample of 960 adults from the Netherlands Twin Register, higher self-rated aggression was also related to lower levels of 3- hydroxybutyrate. These exploratory epidemiologic results warrant further studies on the role of ketone metabolism in aggression.

## Introduction

Aggression is a significant behavioral problem affecting children and adults, with consequences not only for individuals, but families, schools, and communities as well[1]. It is also a complex and heterogenous behavior that has been examined from multiple perspectives, including epidemiologic[2], genetic[3], epigenetic (Van Dongen et al. under review), and biomarkers 4, to name a few. Yet, much variation in aggressive behavior remains to be explained. Some of the mechanisms, pathways, and molecules involved in the biological-level aspects of the behavior are known; however, in both adults and children, there remain biological pathways to discover and biomarkers yet to identify[4–6]. There is a need to generate new hypotheses and approaches to enhance our understanding of aggression and its associated problems.

One such suggested approach is using proton nuclear magnetic resonance (NMR) spectroscopy to find new biomarkers, because it allows for a hypothesis-free way to examine phenotypic associations[7–9]. NMR is not a new technology, but applying the approach to behavioral/psychological problems, including aggression, is gaining interest[4]. The NMR methodology is generally able to produce information on the levels of lipids, amino acids, organic acids, and other small organic molecules. Associations between lipid concentrations and aggression have previously been noted[4,10–13], while other low-molecular-weight molecules might hold more promise for novel findings. Amino acids and ketone bodies (low-molecular-weight molecules categorized in NMR) have been associated with emotional and behavioral problems other than aggression[14–17], and some studies have shown amino acids and similar molecules associated with anger and psychopathy[18], schizophrenia-related violence[13], and childhood aggression[19]. Accordingly, these areas offered a reasonable place to focus our exploratory investigation.

Thus, in this study we combine an NMR approach with the aggression phenotype in efforts to generate new insights into our biological understanding of aggression. We conducted exploratory analyses on the association between aggression and amino acid and ketone body biomarkers, seeking to explore association(s) found in more depth and with an attempt to replicate them in an independent sample. The discovery dataset includes longitudinal aggression measurements in adolescence and biomarker data in young adulthood, while in the replication dataset, biomarker data in adults were measured before the assessment of aggression.

## Materials and Methods

### Discovery Dataset (FinnTwin12)

The FinnTwin12 study is a longitudinal population-based cohort of Finnish twins born in 1983-1987 that was initiated to track behavioral development and health habits from mid-childhood onward[20, 21]. Twins and their families were identified through the Finnish Central Population Registry and initial enrollment of the twins occurred between the ages of 11 and 12. Baseline response rate was 87% (N=5600 twins) and remained high (between 85 and 90199 cl:537) for all data collection waves. Questionnaire data collection waves occurred around ages 11/12, 14, 17, and 22. Data were collected from parents, twins, and teachers at age 11/12 and 14, and from twins at ages 17 and 22.

Additionally, a subset of twins (from 1035 families) was more intensively studied using additional assessments (semi-structured psychiatric interview and questionnaires) in young adulthood (mean age = 22.4, SD = 0.70; n = 1347, response rate 73.0%), as well as a plasma sample, anthropometrics and neuropsychological tests for twins tested in person in young adulthood (n=779).

A blood sample was collected after an overnight fast with the request that the participant abstain from alcohol and tobacco since the night before the sample. Plasma was extracted immediately and stored at −80°C. Samples were processed in one batch in the autumn of 2010 using an automated high-throughput serum NMR metabolomics platform (i.e., the Brainshake/Nightengale platform)[21–23]. The metabolites utilized here were the amino acids (alanine, glutamine, histidine, isoleucine, leucine, phenylalanine, tyrosine, valine) and ketone bodies (acetate, acetoacetate, 3-hydroxybutyrate). All metabolite data were available with units mmol/l. Pregnant women (n=53) and individuals on cholesterol medication (n=1) were excluded from analysis. Thus, the final sample for study was 725 twin individuals.

Ethical approval for all waves of data collection was obtained from the ethical committee of the Helsinki and Uusimaa University Hospital District and Indiana University’s Institutional Review Board. At ages 12 and 14, parents provided consent for the twins, while twins themselves provided written consent for age 17 and 22 collection waves.

### Aggression Measurement

Behavioral and emotional development in FinnTwin12 were measured at ages 12, 14, and 17 using the modified Multidimensional Peer Nomination Inventory(MPNI)[24]. The MPNI is a 37-item questionnaire producing multiple subscales including aggression (6 items; 4 direct aggression items, 2 indirect aggression items). Each question about the child’s behavior (e.g., “Does the child call people names when angry with them?”) has four response choices: ‘not observed in child’, ‘observed sometimes, but not consistently’, ‘definitely observed, but is not prominent’, and ‘clearly observed’. Response choices are scored 0–3, and aggression scores for individual raters (e.g.,teacher rating at age 12) are formed by taking the mean of all items in the aggression subscale (no missing values were allowed). An overall aggression score was created (using all 6 items). Additionally, a composite aggression score (called ‘Aggression Combined’) was used for initial analyses and was formed by calculating the mean of the aggression subscale scores for the parent rating at age 12 and the teachers ratings at ages 12 and 14 combined.

### Covariates

Several covariates, measured at the time of sample collection, were included in the regression model. Body mass index (BMI) was calculated as kg/m^2^ (mean: 23.3, standard deviation (SD): 3.94). Leisure-time physical activity was expressed as metabolic equivalent of task (MET) hours/day based on structured questions considering the intensity, frequency, and duration of activities [25]. Smoking status was categorized as current, former, never/experimenter. Alcohol consumption frequency was categorized as none, once a month or less, 2–4 times per month, 2–3 times per week, 4 or more times per week. Finally, self-rated general health was dichotomized to either poor health (poor + fair) or good health (good + very good).

### Replication Dataset (NTR)

The Netherlands Twin Register (NTR), established in 1987, collects data on twins and other. Data collection is nationwide via questionnaires mailed to parents of young twins (ages 1–12) and to twins themselves at age 14 and beyond. Twins recruited as adolescents and adults and their families are invited to take part in data collection by survey every 2 to 4 years [26]. Multiple survey waves have included the Achenbach System of Empirically Based Assessment (ASEBA) Adult Self-Report (ASR)[27], which includes questions that allow for the calculation of an aggression score. A subgroup of adult participants was invited into the NTR Biobank project [28,29]. As part of this project, they were visited at home where samples were collected between 7:00 and 10:00 after overnight fasting. Participants were asked not to smoke before sampling. Fertile women took part from day 2–4 of their menstrual cycle, or in the pill-free week. During the visit, body composition measurements were taken and information regarding physical health and lifestyle behavior (e.g., smoking and drinking patterns, exercise, medication use) was collected.

Metabolite data, processed using the same Brainshake/Nightingale platform as FT12[23], were assessed in 5204 participants (age range: 18–87 years). We excluded women who were pregnant (n=19) and all individuals over the age of 30 years at the time of sampling (n=3843), to make the FinnTwin12 and NTR samples similar in age composition. The two data collection timepoints (wave 8 and 10) that obtained aggression scores (ASR questionnaire) and were closest to the time of blood sampling were analyzed. ASR aggression scores at wave 8 were available from 819 participants (mean age: 28.1 years, range 18–38 years), and ASR aggression scores from wave 10 from 665 participants (mean age: 32.7 years, range 21–39 years). The aggression score used for the analyses was the score closest to the time of blood sample collection. Of note, aggression scores were generally available closer to the time of sampling for the NTR dataset (mean 3.4 years; range 0–10 years) than the FinnTwin12. There were 267 participants who had aggression score values collected before their blood sample, and these scores were excluded to make the dataset temporally uniform (blood samples first, aggression scores later; this is the opposite of the temporality of the FinnTwin12 sample). Thus, the final sample for study was 960 individuals. For replication, only the metabolite 3-hydroxybutrate (mmol/l) was examined.

NTR Projects were approved by the Central Ethics Committee on Research Involving Human Subjects of the VU University Medical Centre, Amsterdam, an Institutional Review Board certified by the U.S. Office of Human Research Protections (IRB number IRB00002991 under Federalwide Assurance FWA00017598; IRB/institute code NTR 03–180).

### Aggression Measurement

Aggression in the Adult NTR was analyzed from surveys 8 and 10, which both included the ASR, part of the ASEBA for assessing problems and strengths in children and adults, with multiple subscales including aggression [27]. Each aggression question has three response choices which were summed over items, if the number of missing items was not larger than three. For participants with no more than 3 missing values, missing items were imputed. If participants had completed the ASR at both surveys 8 and 10, we took their aggression score closest to the blood sampling (but always after blood sampling).

### Covariates

We aimed to mirror variables and categories as close to the FinnTwin12 dataset as possible. BMI was calculated as kg/m^2^ (from the biobank home visit) (mean: 23.0, SD: 3.43). Leisure-time physical activity (from the same survey wave that the aggression score was taken) was calculated as MET minutes. Smoking status was categorized as current, former, never (from the biobank home visit). Alcohol consumption frequency (from the same survey wave as the aggression score) was categorized as none, once a month or less, 2–4 times per month, 2–3 times per week, 4 or more times per week. Self-rated general health (from the same survey wave that the aggression score was taken) was dichotomized to either poor health (poor + fair) or good health (reasonable + good + very good). Since the NTR metabolite data were processed in different batches, there was a batch variable available for adjustment.

### Analysis

Exploratory analyses in FinnTwin12 involving the available low-molecular-weight metabolites (n=11; 8 amino acids and 3 ketone bodies) utilized the composite aggression score (Aggression Combined) in order to minimize multiple testing and to combine information from different informants, who tend to rate aggression slightly differently [30,31]. Spearman correlations were computed between the untransformed metabolites, and between the metabolites and the Aggression Combined variable. Thereafter, due to their non-normal distribution, all metabolites were rank transformed (generally the best solution for most of the metabolites). Then, linear regression was performed with individual metabolites as the dependent variable and Aggression Combined as the main independent variable, adjusted for age, sex, and BMI. To obtain robust standard errors (i.e., corrected for familial relatedness), the cluster option in Stata was specified [32]. Beta coefficients and 95% confidence intervals (CI) were reported and those reaching significance (p<0.05) were investigated further. After exploratory analyses were performed, we then used an independent sample to see if these were replicated.

To further investigate 3-hydroxybutyrate (the only biomarker that reacheda nominal p<0.05), additional linear regression modeling was employed to establish consistency of the association. In addition to the primary Aggression Combined variable, we examined the separate overall aggression ratings from different raters as well (parents at age 12, teacher(s) at age 12, teacher(s) at age 14, self ratings at ages 14 and 17, and the twin’s co-twin at ages 14 and 17). Initial models designated 3-hydroxybutyrate as the dependent variable and the different overall aggression ratings as the main independent variable, adjusted for age, sex, BMI, and familial relatedness. Further models additionally adjusted for leisure-time physical activity (METs), smoking status, alcohol consumption frequency, and self-rated general health. Sex interactions were also investigated. Supplemental post hoc analysis investigated whether including multiple overall aggression ratings in the same model affected the results.

To obtain further evidence to support or refute the association found in our discovery sample, we analyzed NTR data to test replication of our results. The initial Spearman correlation of the untransformed 3-hydroxybutyrate and the aggression rating was computed. Then, the 3- hydroxybutyrate variable was rank transformed, as in the FinnTwin12 data. Linear regression models were run with 3-hydroxybutyrate as the dependent variable and the aggression rating as the independent variable, adjusted for age, sex, BMI, batch and familial relatedness. Additionally, fully adjusted models further adjusted for leisure-time physical activity (METs), smoking status, alcohol consumption frequency, and self-rated general health. Sex interactions were also investigated to study if the aggression-3-hydroxybutyrate relationsip differed in males and females[33].

Finally, a random effects meta-analysis was conducted on the FinnTwin12 (using the Aggression Combined variable) and NTR fully adjusted models to estimate an overall effect size and test for heterogeneity (I^2^) between the samples. All analyses were performed using Stata version 15.1 or 16.0 (Stata Corporation, College Station, TX, USA).

## Results

### Discovery (FinnTwin12) Results

Spearman correlations of untransformed metabolites indicated generally low to moderate associations, except the high correlations between 3-hydroxybutyrate and acetoacetate (r=0.70)and the branched-chain amino acids (leucine, isoleucine, and valine) with each other (r range=0.62–0.73) Table 1. Spearman correlations between metabolites and the Aggression Combined variable were low, ranging from −0.13 to 0.14.

**Table 1.** FinnTwin12 Spearman correlations of untransformed metabolites and aggression (n=526)

|  | ace | acace | bohbut | ala | gln | his | ile | leu | phe | tyr | val |
| --- | --- | --- | --- | --- | --- | --- | --- | --- | --- | --- | --- |
| acace | 0.11 |  |  |  |  |  |  |  |  |  |  |
| bohbut | 0.15 | 0.70 |  |  |  |  |  |  |  |  |  |
| ala | -0.12 | -0.38 | -0.36 |  |  |  |  |  |  |  |  |
| gln | 0.36 | -0.03 | -0.05 | 0.06 |  |  |  |  |  |  |  |
| his | 0.07 | -0.01 | -0.11 | 0.20 | 0.00 |  |  |  |  |  |  |
| ile | 0.11 | 0.22 | -0.07 | 0.19 | 0.28 | 0.09 |  |  |  |  |  |
| leu | 0.12 | 0.13 | -0.04 | 0.08 | 0.17 | 0.02 | 0.73 |  |  |  |  |
| phe | -0.06 | -0.03 | -0.05 | 0.16 | -0.32 | 0.30 | 0.15 | 0.34 |  |  |  |
| tyr | 0.16 | -0.12 | -0.23 | 0.38 | 0.39 | 0.20 | 0.42 | 0.30 | 0.15 |  |  |
| val | 0.20 | 0.16 | -0.02 | 0.07 | 0.37 | 0.05 | 0.68 | 0.62 | 0.03 | 0.55 |  |
| AggComb<sup>a</sup> | -0.04 | 0.04 | -0.13 | 0.04 | 0.09 | 0.07 | 0.14 | 0.10 | -0.06 | 0.13 | 0.11 |

Abbreviations: ace=acetate; acace=acetoacetate; AggComb=Aggression Combined; bohbut=3-hydroxybutyrate; ala=alanine; gln=glutamine; his=histidine; ile=isoleucine; leu=leucine; phe=phenylalanine; tyr=tyrosine; val=valine
<sup>a</sup>aggression is 'Aggression Combined' variable (which is the mean of parent-12, teacher-12, and teacher-14 aggression ratings)

Next, linear regression models with rank-transformed metabolites as the dependent variable and Aggression Combined as the main independent variable, also adjusted for age, sex, BMI, and familial relatedness, revealed that only 3-hydroxybutyrate was significantly associated with aggression (Figure 1; Supplemental Table 1). The association was negative, with lower 3- hydroxybutyrate levels being associated with higher aggression levels.

**Fig 1.**
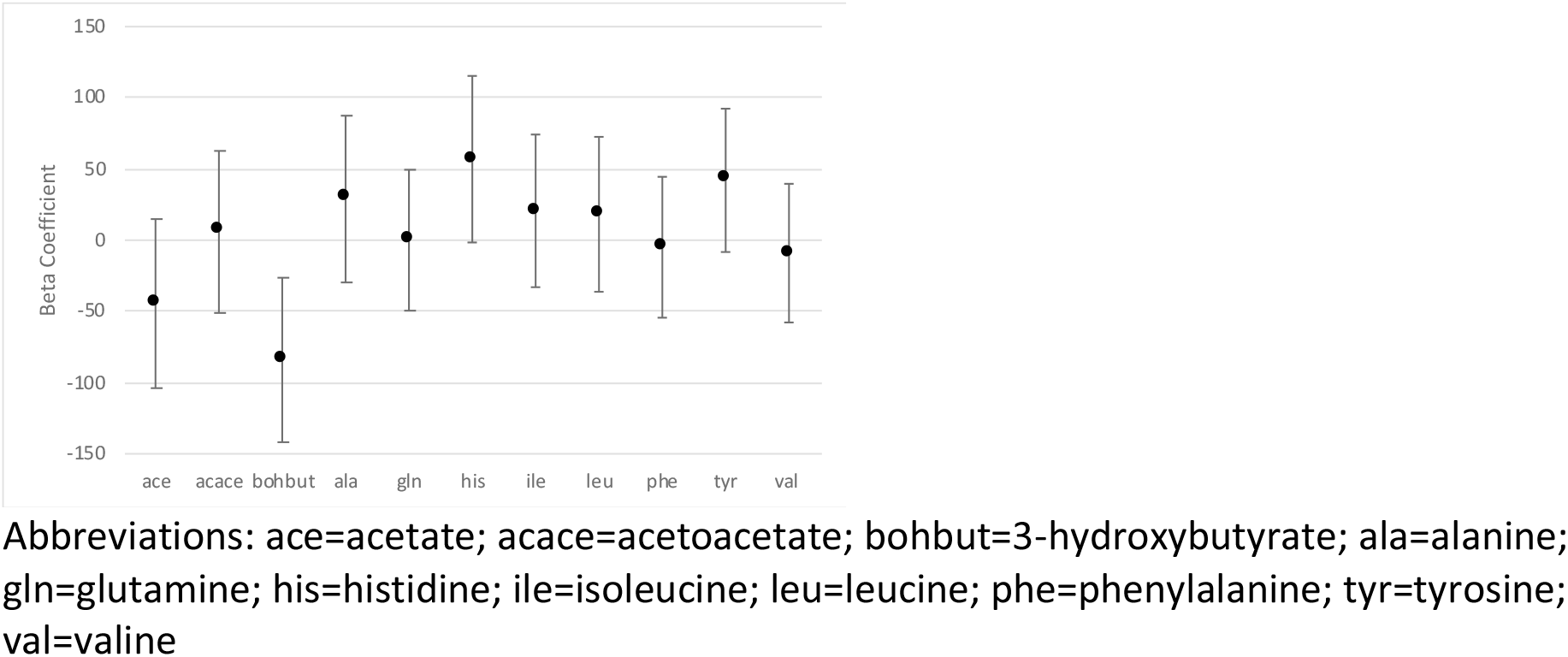
FinnTwin12 beta coefficients (and 95% confidence intervals) of Aggression Combined independent variable in linear regression models with metabolites as dependent variables, adjusted for age, sex, body mass index, and familial relatedness.

In linear regression models further exploring 3-hydroxybutyrate, which showed the strongest association with aggression, the basic model results indicated that the teacher ratings of aggression at age 12 and self ratings at age 14 showed the strongest associations (and were also statistically significant), though nearly all ratings supported the trend of negative association of 3- hydroxybutyrate with aggression (Figure 2;Supplemental Table 2). In fully adjusted models (basic model plus MET, smoking status, alcohol consumption frequency, and self-rated general health), the negative associations remained, with only small attenuation to the effect sizes (Table 2). No sex interactions were detected (p-value range=0.10–0.84; Supplemental Table 3).

**Fig 2.**
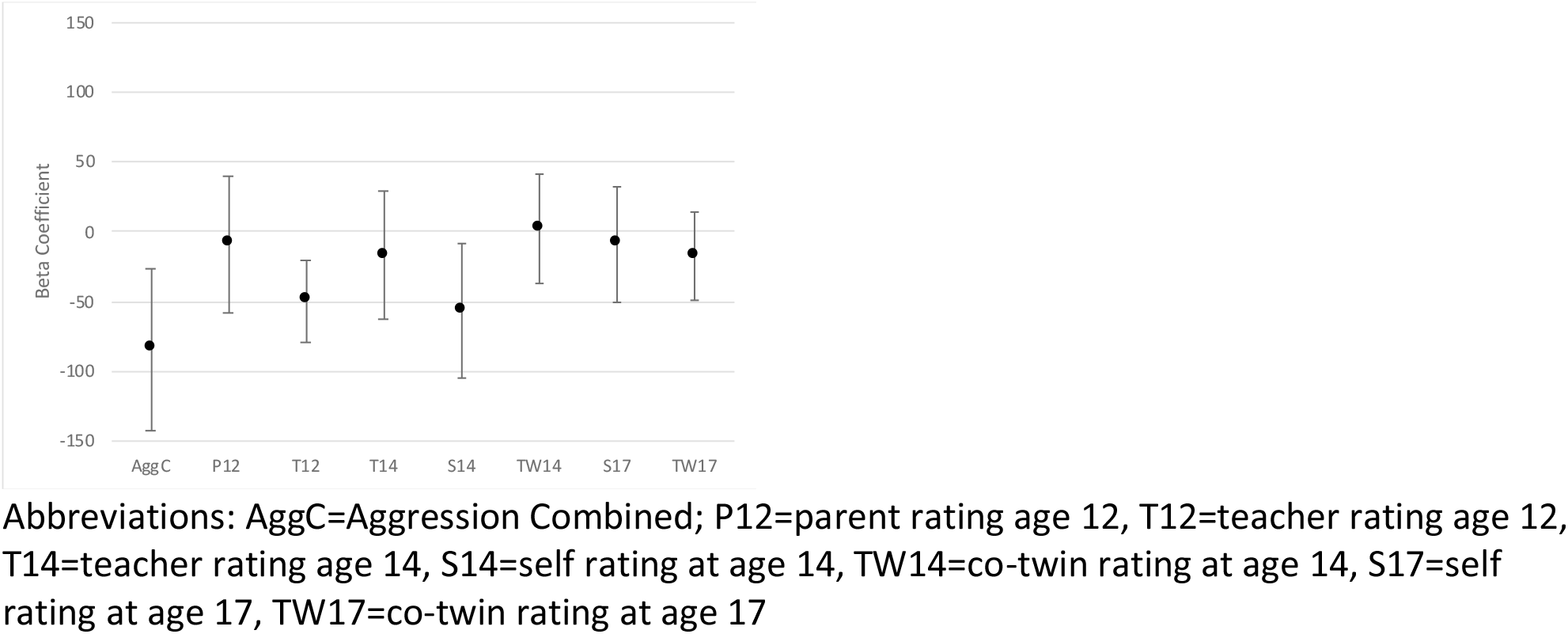
FinnTwin12 beta coefficients (and 95% confidence intervals) of aggression ratings from different raters as (separate) independent variables in linear regression models with rank-transformed 3-hydroxybutyrate as the dependent variable, adjusted for age, sex, body mass index, and familial relatedness.

**Table 2.**
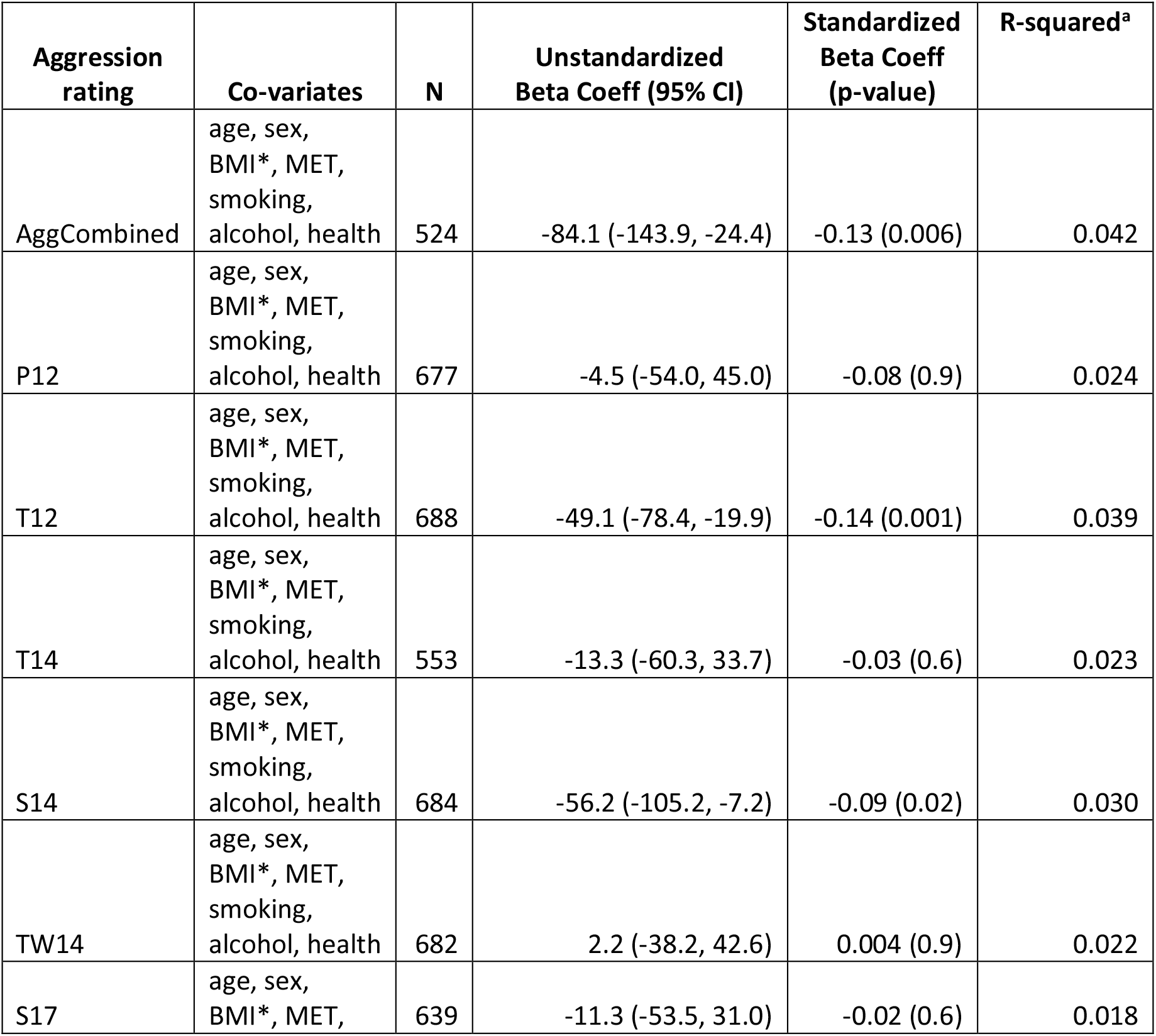
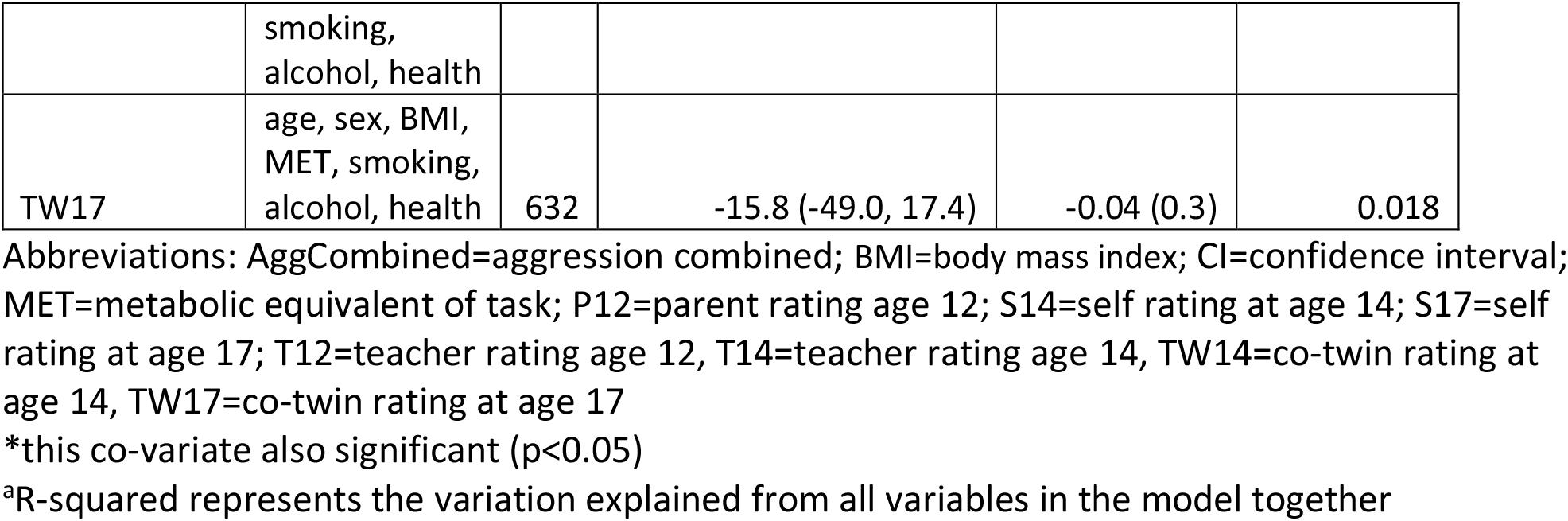
FinnTwin12 linear regression models with 3-hydroxybutyrate as the dependent variable, beta coefficients of aggression ratings from different raters as independent variables in separatemodels, adjusted for age, sex, BMI, MET, smoking status, alcohol consumption frequency, self-ratedgeneral health, and familial relatedness.

Supplemental results indicated that the separate aggression scores by different raters at different twin ages were only weakly to moderately correlated (r range: 0.14–0.47). Thus, multiple aggression ratings were added into the same model as independent variables, adjusted for age, sex, BMI, and familial relatedness. The tested combinations included the three aggression scores that made up the Aggression Combined variable (parent age 12, teacher age 12, teacher age 14), both age 12 ratings, all three age 14 ratings, and both age 17 ratings, as well as the two ratings that were consistently significant (teacher age 12 and self age 14). Interestingly, in the model with both teacher age 12 and self age 14 ratings, both were statistically significantly negatively associated with 3-hydroxybutyrate (Supplemental Table 4).

### Replication (NTR) Results

The Spearman correlation between the aggression score and untransformed 3-hydroxybutyrate was −0.07 (p=0.045) in the replication dataset. The initial linear regression model with rank-transformed 3-hydroxybutyrate as the dependent variable and aggression as the main independent variable, adjusted for age, sex, BMI, batch and familial relatedness, showed the same negative association trend (p=0.138) as the FinnTwin12 models Table 3. There was no sex interaction detected in the initial model (p=0.246). However, in the fully adjusted model there was a significant sex interaction (p=0.015). Regarding the fully adjusted models, the negative association is present for males only; it became slightly positive in direction for females, albeit with wide confidence intervals. Table 3 displays results for the sexes pooled and males and females modeled separately.

**Table 3.**
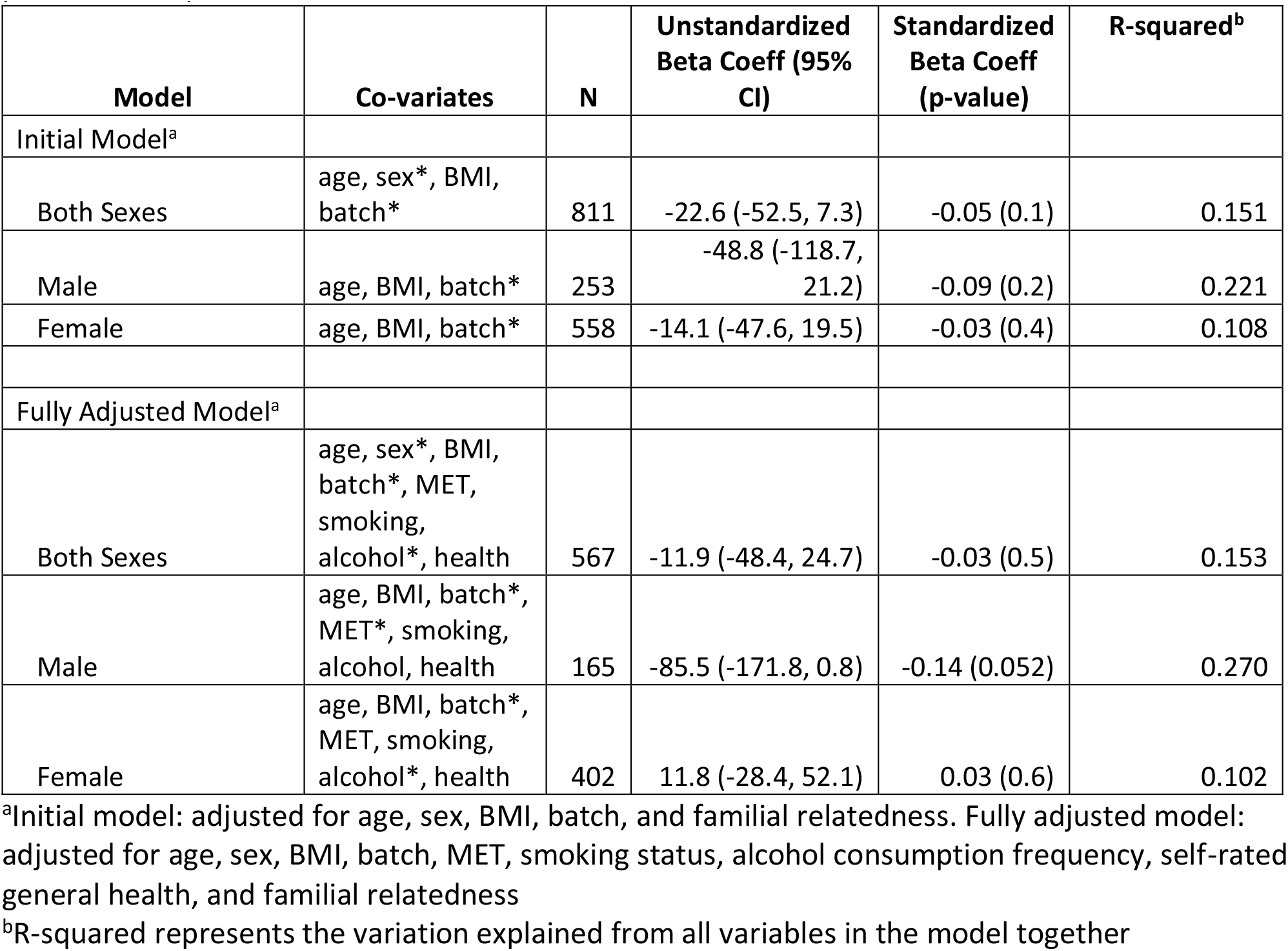
NTR linear regression models (initial and fully adjusted^a^) with 3-hydroxybutyrate as the dependent variable, beta coefficients of aggression rating as independent variable, with sexes pooled and separated

### Meta-analysis

The random effects meta-analysis calculation comparing the FinnTwin12 and NTR fully adjusted sex-pooled models indicated an overall effect size of −295 cl:59344.0 (–114.4, 26.4)(Supplemental Table 5). The test for heterogeneity revealed moderate to high heterogeneity between the two datasets (I^2^=75.7%). Meta-analysis results for the four fully adjusted sex-separated models from FinnTwin12 and NTR are also available in Supplemental Table 5.

### Discussion

In this study, we showed an association between aggression and a ketone body biomarker, 3- hydroxybutyrate. We initially tested 11 potential NMR-processed low-molecular-weight biomarkers for association with a composite aggression score (included parent and teacher ratings of aggression of children at ages 12 and 14). 3-hydroxybutyrate was negatively associated with aggression and was the only nominally statistically significant biomarker. Upon further analysis, in both basic (age, sex, and BMI) and fully adjusted (age, sex, BMI, MET, smoking status, alcohol consumption frequency, self-rated general health) models, eight different aggression ratings (from different raters and ages) were tested. They showed a consistent pattern of negative association, as seven out of eight showed the negative association with 3-hydroxybutyrate, with three of them being statistically significant. When including both teacher at age 12 and self at age 14 aggression ratings in the same model, both aggression ratings were significantly associated with 3-hydroxybutyrate, showing that an external rater (teacher) and an adolescent’s self-rating contribute separately to this association; this phenomenon has been seen in previous studies (Whipp 2019) and relates to the known low interrater correlations among behavioral phenotypes [30]. Importantly, we were able to assess this biomarker-aggression association in a large and independent sample, the NTR. In both basic and fully adjusted models, the negative association of 3-hydroxybutyrate and aggression was found, but statistical significance was not reached until the sexes were separated in the fully adjusted model (as indicated by the significant sex interaction). Finally, meta-analysis indicated that while there was moderately high heterogeneity between the samples, there remained an overall effect size showing the negative association: low 3- hydroxybutyrate levels with high aggression levels.

To our knowledge, this is the first time ketone bodies, and 3-hydroxybutyrate specifically, have been associated with aggression. In a urinary metabolite study with aggression in the NTR, no significant associations were found, but several near-significant results pointed to energy metabolism as a potential group of metabolites warranting further investigation[19]. Additionally, there are two recent studies showing a link between 3-hydroxybutyrate and depression, one in mice that was negatively correlated [16] and one in humans that was positively correlated [17]. We know from previous studies that depression and aggression often co-occur [34,35], and the concept of a ‘p factor’ that potentially underlies all psychopathology has been gaining traction [36]. However, our finding of a negative aggression-3-hydroxybutyrate relationship is in contrast with earlier reported positive depression-3-hydroxybutyrate relationships and warrants studying these traits separately in relation to metbolites.

In evaluating the biological plausibility of this association, we can consider generally what we know about this compound. During periods of exercise or fasting (low availability of glucose as an energy source), ketone bodies become involved in an alternative energy pathway [37]. 3- hydroxybutyrate, in particular, has been shown to be not only involved in the alternative energy pathway, but it also has important cell signaling and regulatory functions, including pathways involved in reducing oxidative stress and inflammation (both linked to aggression: [5,38] and epigenetic modifications [17,39]. We also know that while 3-hydroxybutyrate is generally produced in the liver, it can cross the blood-brain barrier and act in brain tissue [40,41]. Finally, although there appears complex biological mechanisms at play, ketogenic diets (which increase ketone body levels) have been popular treatments in neurologic disorders, such as epilepsy, for decades [42]. The field of psychiatry has recently become more interested in the potential benefits of this diet, including in attention-deficit disorders and autism [43], both of which can co-occur with aggression, however, aggression and the ketogenic diet has yet to be considered.

This study suggesting a new biomarker of aggression has important strengths in its large sample size, multiple ratings of aggression across stages of development, and the direction of association replicated in an independent sample. There are also limitations. First, we did not have aggression measured at the same time as the biological sample, in either dataset. However, it is known that aggression is a relatively stable trait, especially in aggressive males. Although aggression levels in childhood and adolescence are generally higher than in adulthood, aggression levels are relatively stable in their rank order among similar aged peers [44,45]. There is longitudinal continuity in aggression from adolescence to adulthood [46], which is the development period covered in the present study. Additionally, a fairly recent study with similar temporal data collection limitations, using childhood aggression measures (as developmental trajectories) and biological samples in young adulthood, was able to identify cytokines (a measure of inflammation) as a novel group of biomarkers for aggression[5]. Regarding biomarker stability, while we do see high heritability in the energy metabolism biomarkers (53.2% for 3-hydroxybutyrate)[47], it would also be important in future studies to have repeated biological samples to confirm the stability of this biomarker over time, concurrent with the stability of aggression measures. However, repeated blood samples from children may be challenging to obtain, and thus this work might need to be done in young adults first, or using less invasive samples such as urine or saliva, which might produce different results than plasma (the recent urine biomarker study on NTR children did not show any significant findings with 3-hydroxybutyric acid and aggression)[19].

In conclusion, this exploratory study using NMR-processed metabolites in plasma samples has revealed a potential new biomarker of aggression: 3-hydroxybutyrate, a ketone body related to an alternative energy pathway. We observed a negative association — high aggression levels with low ketone bodies levels — in our Finnish dataset, which was suggestively replicated in our independent Dutch dataset. There is biological plausibility for associating this ketone body aggression, as well as the ability to potentially influence the biological levels of 3-hydroxybutyrate via diet modificiation. Thus, it is well-warranted to further investigate this epidemiological association at the biological level, hopefully shedding more light on the complexities of the aggression phenotype.

## Data Availability

The datasets generated during and/or analysed during the current study are not publicly available due to participant confidentiality, but are available from the corresponding author on reasonable request.

## Acknowledgements

FinnTwin12 wishes to thank all participating twins, their parents and teachers. Data collection in FinnTwin12 was supported by NIAAA12502, AA-00145, AA-08315 and the Academy of Finland (grants 100499, 205585, 118555, 141054 and 264146 to JK). JK has been supported by the Academy of Finland (grants 308248 and 312073).

NTR warmly thanks all twins and family members for their participation. NTR data collection and analyses were supported by the Netherlands Organization for Scientific Research: Netherlands Twin Registry Repository: researching the interplay between genome and environment (480-15- 001/674); the European Union Seventh Framework Program (FP7/2007-2013): ACTION Consortium (Aggression in Children: Unravelling gene-environment interplay to inform Treatment and InterventiON strategies; grant number 602768) and Biobanking and Biomolecular Resources Research Infrastructure Netherlands (BBMRI –NL: 184.021.007 and 184.033.111).

## Author Contributions

AMW, DIB, and JK made substantial contributions to the conception/design of the study;DIB, JK, LP, MB, and RJR made substantial contributions to the acquisition of the data;AMW, EV, JK, LHB, RP, RSLL, and TK made substantial contributions to the analysis of the data; AMW, DIB, EV, FAH, JK, LHB, RP, RSLL, and TK made substantial contributions to the interpretations of the data;

AMW drafted the manuscript; and all authors made substantial contributions to the revision of the manuscript draft

